# Comparative B cell and antibody responses induced by adenoviral vectored and mRNA vaccines against COVID-19

**DOI:** 10.1101/2023.06.02.23290871

**Authors:** Yi Liu, Stephany Sánchez-Ovando, Louise Carolan, Leslie Dowson, Arseniy Khvorov, Jessica Hadiprodjo, Yeu Yang Tseng, Catherine Delahunty, Ameneh Khatami, Marion Macnish, Sonia Dougherty, Michelle Hagenauer, Kathryn E. Riley, Ajay Jadhav, Joanne Harvey, Marti Kaiser, Suja Mathew, David Hodgson, Vivian Leung, Kanta Subbarao, Allen C. Cheng, Kristine Macartney, Archana Koirala, Helen Marshall, Julia Clark, Christopher C. Blyth, Peter Wark, Adam J. Kucharski, Sheena G. Sullivan, Annette Fox

## Abstract

Both vector and mRNA vaccines were an important part of the response to the COVID-19 pandemic and may be required in future outbreaks and pandemics. However, adenoviral vectored (AdV) vaccines may be less immunogenic than mRNA vaccines against SARS-CoV-2. We assessed anti-spike and anti-vector immunity among infection-naïve Health Care Workers (HCW) following two doses of AdV (AZD1222) versus mRNA (BNT162b2) vaccine. 183 AdV and 274 mRNA vaccinees enrolled between April and October 2021. Median ages were 42 and 39 years, respectively. Blood was collected at least once, 10-48 days after vaccine dose 2. Surrogate virus neutralization test (sVNT) and spike binding antibody titres were a median of 4.2 and 2.2 times lower, respectively, for AdV compared to mRNA vaccinees (p<0.001). Median percentages of memory B cells that recognized fluorescent-tagged spike and RBD were 2.9 and 8.3 times lower, respectively for AdV compared to mRNA vaccinees. Titres of IgG reactive with human Adenovirus type 5 hexon protein rose a median of 2.2-fold after AdV vaccination but were not correlated with anti-spike antibody titres. Together the results show that mRNA induced substantially more sVNT antibody than AdV vaccine due to greater B cell expansion and targeting of the RBD. Pre-existing AdV vector cross-reactive antibodies were boosted following AdV vaccination but had no detectable effect on immunogenicity.

**Key points:**

- mRNA SARS-CoV-2 vaccine induced higher surrogate neutralizing antibody titres than adenoviral vaccine
- mRNA vaccine induced a more potent, RBD-targeted B cell response than AdV vaccine
- Adenoviral vaccine boosted antibodies against human Adenovirus, but titres don’t correlate with anti-spike titres

## Background

The COVID-19 pandemic accelerated the development and wide-spread use of vaccines that encode SARS-CoV-2 spike protein as mRNA or as DNA delivered in a viral vector ^1^. The Adenovirus-vectored (AdV) SARS-CoV-2 vaccine, AZD1222 (Vaxzevria®, AstraZeneca) contains a replication defective chimpanzee adenovirus vector (ChAdOx1) expressing a codon-optimised sequence for the full-length spike of SARS-CoV-2 ^2^. The BNT162b2 (Comirnaty®, Pfizer) vaccine contains nucleoside-modified RNA encoding membrane-anchored full-length spike of SARS-CoV-2 formulated within lipid nanoparticles^3^. The encoded protein is stabilized in the prefusion conformation by substituting two amino acids in the C-terminal S2 fusion machinery to prolines (K986P and V987P) ^4, 5^.

Clinical trials of AZD1222 report efficacy of 74-90% ^6–8^, and trials of BNT162b2 report efficacy of 86-100% ^3, 9^. Receptor binding domain (RBD) reactive and neutralizing antibody (nAb) titres correlate with protection and account for around two thirds of vaccine efficacy ^10, 11^. While early trials indicated that the vast majority of healthy adults develop nAbs after two doses either vaccine ^2, 12, 13^, an immunogenicity trial in older adults found that antibody levels were lower for AdV compared to mRNA vaccinees ^14^. Similarly, a number of small field studies indicate that anti-spike antibody levels are low for AdV compared to mRNA vaccinees ^15–18^.

Vector vaccines may be required in the event of future outbreaks and pandemics, and are the most advanced vaccines against Ebola. For COVID-19, these vaccines were less sensitive to temperature making them more appropriate for distribution in resource-limited settings compared with mRNA vaccines which required freezing. Therefore, it is important to further investigate if and why immunogenicity differs between AdV and mRNA vaccines, in particular whether anti-vector immunity may compromise AdV immunogenicity, as this technology may be needed in the future.

In Australia, Comirnaty® mRNA and Vaxzevria® AdV vaccines were the first COVID-19 vaccines approved for use and were provided free of charge. Rollout was staged commencing with health and aged care workers, those working in quarantine facilities, and high-risk groups secondary to age or comorbidities. Two-dose regimens with intervals of 21d for mRNA and 90d for AdV were recommended. Australian governments implemented strict control strategies that minimized SARS-CoV-2 transmission until high vaccination coverage was achieved ^19^. Therefore, health care workers (HCWs) were rarely infected prior to receiving their second dose of vaccine, providing an opportunity to compare the immunogenicity of mRNA versus AdV vaccines without interference from pre-existing immunity induced by infection.

In this study, we investigated the immunogenicity of AZD1222 AdV versus BNT162b2 mRNA vaccines in healthcare workers at 6 Australian hospitals. Sera collected from all participants around 14 days after dose 2 of vaccination were used to compare surrogate virus neutralization test (sVNT) antibody titres. Additional analyses were conducted to explore whether differences in immunogenicity may be associated with the extent to which B cells and antibodies target the RBD of spike. Finally, we also explored whether there could be any effect of pre-existing human adenovirus reactive antibodies.

## Methods

### Study design

In April 2020, a prospective cohort study (ClinicalTrials.gov Identifier: NCT05110911) was established to investigate influenza vaccine immunogenicity among Health Care Workers (HCWs) at six health services across Australia (Alfred Hospital, Victoria; Children’s Hospital Westmead and John Hunter Hospital, New South Wales; Perth Children’s Hospital, Western Australia; Queensland Children’s Hospital; and the Women’s and Children’s Hospital Adelaide, South Australia). Commencing April 2021, the study pivoted to enable follow-up of COVID-19 vaccination.

HCWs, including medical, nursing, and allied health staff, students and volunteers aged 18Y to 60Y, were recruited at each hospital’s staff influenza vaccination clinic or responded to recruitment advertising. Those on immunosuppressive treatment (including systemic corticosteroids) within the past 6 months, and contraindicated for vaccination were excluded. Enrolled participants provided a 9ml blood sample for serum collection ∼14 days after their second dose of COVID-19 vaccine (suggested range 10-21 days). Pre-vaccination blood samples were collected from participants who enrolled prior to receiving a first dose of vaccine. Alternately, samples collected in 2020 were used for participants who enrolled in the influenza vaccination cohort at that time. End of year sera, collected October through November 2021, were also assessed. A subset of participants provided additional blood samples for peripheral blood mononuclear cells (PBMC) recovery on day 0 if enrolled prior to receiving their first vaccine dose and ∼ 7 and 14 days after vaccination. Participants completed weekly symptoms diaries to monitor for acute respiratory illnesses (ARI), defined as one or more of fever ≥37.8°C, headache, body aches, cough, sore throat, runny nose, or sputum. In the event of an ARI, they were asked to provide a self-collected nose/throat swab for PCR testing; however, public health measures implemented during the COVID-19 pandemic required participants to attend government testing clinics, and many did not self-swab.

The study protocol and protocol addendums for follow-up of COVID-19 vaccinations and SARS-CoV-2 infections were approved by The Royal Melbourne Hospital Human Research Ethics Committee (HREC/54245/MH-2019).

### Surrogate Virus Neutralization Test (sVNT) assay

The SARS-CoV-2 sVNT assay described by Tan et al ^21^ was adapted to utilize commercially available SARS-CoV-2 spike receptor binding domain (RBD) protein (SinoBiological, 40592-V27H-B) representative of the ancestral strain (YP_009724390.1). Sera were serially diluted 3-fold from 1:10 to 1:21870 for testing. GraphPad Prism version 9.5.1 for Windows (GraphPad Software, California USA) was used to fit sigmoidal curves of OD450 values against log10 serum dilutions and to interpolate 50% inhibition titres. Sera that had no detectable inhibition at the lowest dilution were assigned a value of 1. Full details are provided in Supplementary Materials.

### Spike binding antibody ELISA

Spike binding antibodies were detected in serially diluted sera using WANTAI SARS-CoV-2 Ab ELISA (Wantai Biological Pharmacy Enterprise Co., Ltd. Beijing 102206, China) according to the manufacturer’s instructions. GraphPad Prism was used to fit sigmoidal curves of OD450 values against log10 serum dilutions and to interpolate 50% antibody binding titres.

### SARS-CoV-2 spike- and RBD-reactive B cell analysis

PBMCs were recovered using Lymphoprep (STEMCELL Technologies, Vancouver, Canada) and LEUCOSEP tubes (Greiner); cryopreserved in FCS containing 10% DMSO; and thawed into RPMI containing DENARASE (cLEcta, Leipzig Germany). Biotinylated spike and RBD were labelled with Streptavidin-fluorochromes (SA-F) using a ratio of 4:1 for SA-F:protein (Supplementary Table 1). PBMCs were incubated with fluorescent-labelled recombinant spike and RBD proteins and with a cocktail of mAbs to detect activated memory B cells (Supplementary Table1). Full details are provided in Supplementary Materials.

### Adenovirus 5 hexon binding IgG ELISA

Human adenovirus type 5 (Ad5) is endemic in humans ^20^. Hexon is a capsid protein targetted by Ad5 neutralizing antibodies ^20^, and is 77% identical between Human Ad5 (GenBank: AAO24091.1) and Chimpanzee AdY25 (NCBI Reference Sequence: YP_006272963.1). Ad5 hexon binding antibody ELISA was performed to determine if titres are boosted by AdV vaccination, indicating that antibodies cross-recognize ChAdOx1 hexon, and if titres correlate with anti-SARS-CoV-2 spike antibodies. Plates were coated with hexon protein overnight, washed and blocked, then incubated with 3-fold dilutions of sera (1:10 to 1:21870). Horseradish peroxidase conjugated anti-human IgG was used to detect bound IgG. Titres were interpolated from sigmoidal curves as described above. Full details are provided in Supplementary Materials.

### Statistical Analysis

Data were summarized as medians and interquartile ranges and compared using Wilcoxon ranked summed tests for continuous variables or Fisher’s exact test for categorical variables, using the stats package in R. Continuous variables were log or square-root transformed to perform correlations using Spearman’s test or multivariate analyses using linear models. All statistical tests were performed in R version 4.2.2. Spearman’s correlation coefficients and associated p-values were obtained from the ggplot package.

## Results

### Participant characteristics

Four-hundred and eighty-six HCWs participated in the COVID-19 vaccination study. Twenty-six were excluded because samples were collected less than 10 or more than 50 days after dose 2 of vaccine. Three additional participants were excluded, two who were infected prior to vaccination, and one who was vaccinated with AdV followed by mRNA vaccine. The final sample included 457 HCWs, of whom 183 received AdV vaccine and 274 mRNA vaccine. Demographic and vaccination details are shown in Table 1. First vaccine doses were administered from 25 February to 16 September 2021; second doses from 18 March to 13 October 2021; and third doses from 9 November 2021. Most (84% in both groups) participants were female. AdV vaccinees were slightly older and had blood collected a median of one day earlier after their second vaccine dose than mRNA vaccinees. The median interval between dose 1 and 2 was 84 days for AdV vaccinees versus 21 days for mRNA vaccinees.

**Table 1.** Demographic and immunization parameters of participants who received AdV versus mRNA vaccine

| **Characteristic** | **Overall,**<br>**N = 457 <sup>a</sup>** | **AdV,**<br>**N = 183** | **mRNA,**<br>**N = 274** | **p-**<br>**value <sup>b</sup>** |
| --- | --- | --- | --- | --- |
| Female | 383 (84%) | 153 (84%) | 230 (84%) | 0.9 |
| Sex unknown | 1 | 0 | 1 |  |
| Age (years) | 40 (32, 49) | 42 (31, 51) | 39 (32, 48) | 0.044 |
| Body Mass Index | 25 (22, 29) | 25 (22, 30) | 25 (22, 28) | 0.064 |
| Presence of any health conditions <sup>c</sup> | 67 (15%) | 35 (19%) | 32 (12%) | 0.031 |
| Days between dose 1 and 2 | 27 (21, 84) | 84 (84, 86) | 21 (21, 24) | <0.001 |
| Days between doses unknown | 1 | 0 | 1 |  |
| Days from dose 2 to testing | 16 (14, 20) | 15 (14, 19) | 16 (14, 21) | <0.001 |

**a:** n (%); Median (interquartile range)
**b:** Fisher's exact test; Wilcoxon rank sum test
**c:** Health conditions of interest included renal disease, chronic respiratory conditions, haematological disorders, chronic neurological conditions, immunocompromising conditions not requiring suppressants, diabetes or other metabolic disorder, pregnancy and smoking.

The first breakthrough infection was detected on December 16, 2021. We could not compare the effectiveness of AdV versus mRNA vaccines or correlate antibody titres with protection, as initially planned, because 123/183 (67%) of AdV vaccinees received an mRNA vaccine booster (dose 3) prior to infection.

### Surrogate virus neutralizing test (sVNT) antibody titres

Median sVNT antibody titres using ancestral strain RBD were 4.2 times higher for mRNA than for AdV vaccine recipients (Figure 1b). Based on interpolation from a standard curve, median sVNT antibody Units per ml were 1362 and 3692 for AdV and mRNA vaccinees, respectively. Thirteen (7.1%) AdV vaccinees had undetectable post-vaccination sVNT antibody titres whereas all mRNA vaccinees had detectable titres. There was a modest decline in sVNT titres with age among mRNA vaccines and male AdV recipients, but not among female AdV vaccinees (Figure 1c). Among mRNA but not AdV vaccinees there was also strong evidence of a modest increase in sVNT antibody titre with increasing interval between dose 1 and 2 (Figure 1d) and a small decrease with increasing time to sample collection (Supplementary Figure 1). sVNT antibody titres remained higher among mRNA compared to AdV vaccinees after adjusting for age (Supplementary Table 2).

**Figure 1.**
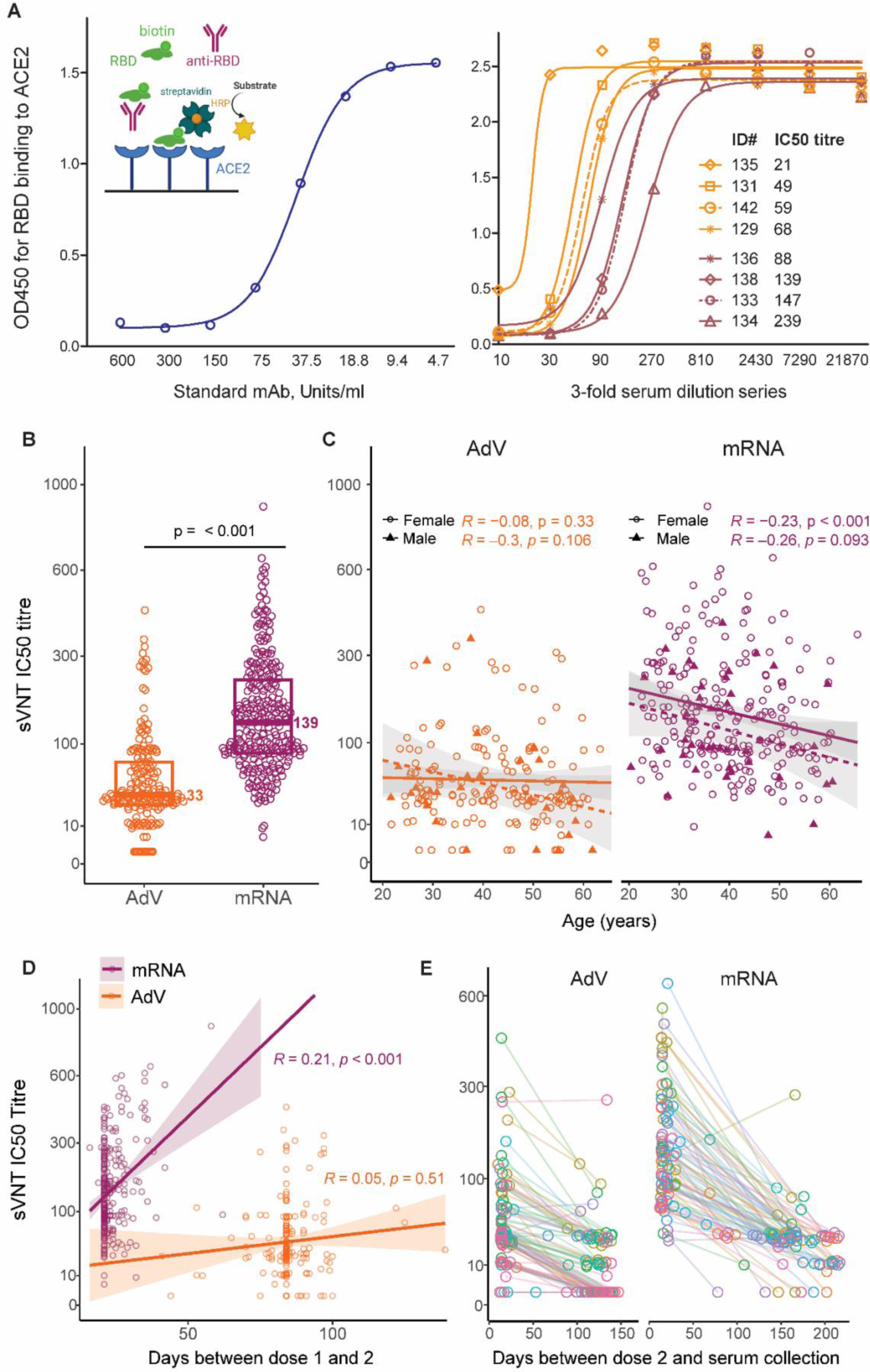
Surrogate virus neutralization test (sVNT) antibody titres by vaccine type, age, sex and intervals between vaccine doses and vaccination and serum collection. (A) assay schematic and mAb standard curve (left panel); raw data for one assay plate (right panel). (B) Titres of sera collected from 183 AdV and 273 mRNA vaccinees around 14 days after their second dose. Symbols represent a single measurement for each individual. Box plots represent medians as bars, with values adjacent, and interquartile ranges as boxes. P-values are shown for comparison of AdV and mRNA vaccinees using Wilcoxon rank-sum test. (C) sVNT antibody titres by participant age and sex. Spearman’s correlation coefficient R and associated p-values are shown. (D) sVNT antibody titres by days between dose 1 and 2 of mRNA vaccine. Spearman’s correlation coefficient R and p-values are shown. (E) sVNT antibody titre decay assessed for a subset of 92 AdV and 93 mRNA vaccinees. Each line and paired symbol represents an individual participant.

436/457 participants had end of year sera collected after their post-COVID-19 vaccination blood draw and before receiving a 3^rd^ dose of vaccine. 93/177 sera from AdV and 93/259 sera from mRNA vaccinees were randomly selected to assess sVNT titre decay. Three AdV and four mRNA vaccinees were excluded from analysis of decay because they had a titre rise exceeding two-fold. Characteristics of included participants are shown in Supplementary Table 3. sVNT titres dropped by a median of 4-fold over a median of 125 days (∼ 3.2 fold/100 days) among AdV vaccinees and 5.1-fold over a median of 162 days among mRNA vaccinees (∼3.1 fold/100 days) (Figure 1e, Supplementary Table 3). Notably, 46% of AdV vaccinees had undetectable sVNT titres by the end of the season time-point compared to only 8% of mRNA vaccinees (Supplementary Table 3).

### Spike binding antibody titres

Ancestral strain spike binding antibody titres were assessed for a randomly selected subset of 96 AdV and mRNA vaccinees. Median 50% spike binding titres were 2.2-fold higher for mRNA vaccinees than for AdV vaccinees (Figure 2a). Spike binding IgG titres rose more steeply against sVNT antibody titres for AdV compared to mRNA vaccinees (Figure 2b).

**Figure 2.**
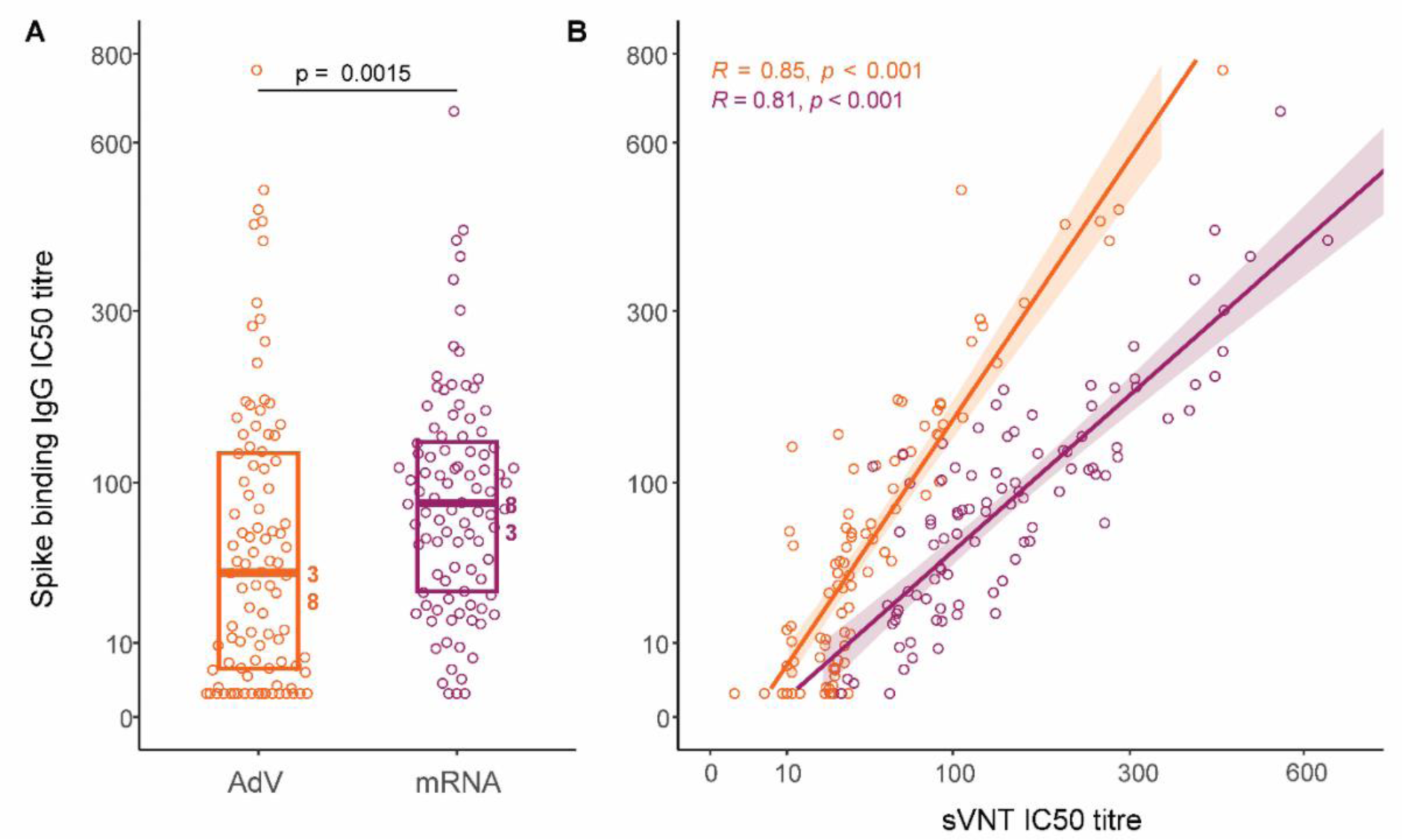
Spike binding IgG titre by vaccine type and by sVNT IC50 titre. (A) Spike binding antibody titres of sera collected from 96 each of AdV (orange) and mRNA vaccines (maroon) around 14 days after their second vaccine dose. Symbols represent a single measurement for each individual. Box plots represent medians as bars, with values adjacent, and interquartile ranges as boxes. P-values from the Wilcoxon rank-sum test for comparison of vaccine types are shown. (B) Correlation between sVNT antibody titre and spike binding IgG titre for each vaccine type. Spearman’s correlation coefficient R and associated p-values are shown.

### SARS-CoV-2 spike- and RBD-reactive memory B cell frequencies

Forty-two HCWs provided additional blood samples for PBMC isolation around day 7 and/or 14 after their second dose of vaccine. Eight received AdV vaccine and had a median age of 54 years. 34 received mRNA vaccine, and were younger, with a median age of 40 years (p = 0.0062). Spike^+^ cells were detected among CD71^+^ memory B cells after vaccination (Figure 3a). Some spike^+^ B cells recognized RBD and were CD20^-^CD38^+^ plasmablasts (Figure 3a). Consequently, spike^+^ B cells frequencies correlated with sVNT antibody titres (Figure 2b).

**Figure 3.**
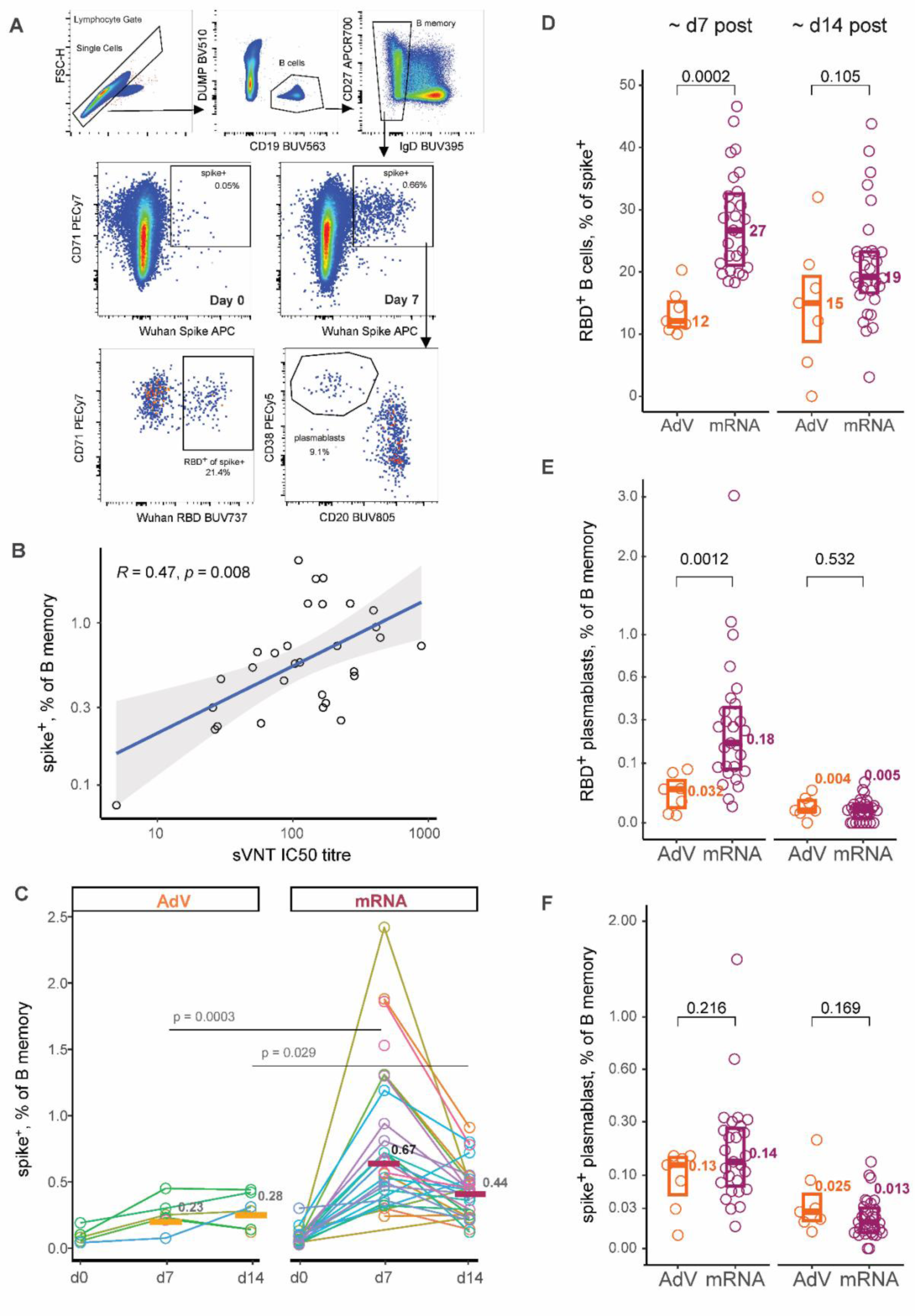
Frequencies of B cells reactive with SARS-CoV-2spike and spike receptor binding domain (RBD) by vaccine type. (A) SARS-CoV-2 spike-reactive B cells were enumerated as a percentage CD27^+^ IgD^-^ memory B cells, then further gated to determine percentages that recognized RBD, or Omicron strain spike or RBD. Data are shown for one mRNA vaccinee. (B) Correlation of spike-reactive B cell frequencies on day 7 with sVNT antibody titres on day 14. Spearman’s correlation coefficient R and associated p-values are shown. (C) Percentages of memory B cells reactive with SARS-CoV-2 spike on day 0 and approximately 7 and 14 days after dose 2 are shown for all individuals assessed. (D) Percentages of spike-reactive B cells that recognize RBD. (E) Percentages of memory B cells that recognize RBD and are CD38^hi^, CD20^-^ plasmablasts. (F) Percentages of memory B cells that recognize spike and are CD38^hi^, CD20^-^ plasmablasts. All boxplots represent medians as bars, with values adjacent, and interquartile ranges as boxes. P values from the Wilcoxon rank-sum test are shown for comparison of vaccine types.

Frequencies of antigen specific B cells, but not of total B cells or B memory cells, differed between participants who received mRNA compared to AdV vaccine on day 7 (Figure 3c, Table 2). Median percentages spike^+^ and RBD^+^ B cells on ∼ day 7 were 2.9 and 8.3 times higher for participants who received mRNA vaccine (Figure 3c, Table 2). Trends were maintained after adjusting for age (Supplementary Table 4). Spike^+^ B cell frequencies declined with increasing days between vaccination and sample collection among mRNA, but not AdV vaccinees (Supplementary Figure 2). Nevertheless, spike^+^ B cell frequencies remained significantly higher for mRNA than for AdV vaccinees on ∼day 14 (Figure 3c). The fraction of spike^+^ B cells that targeted RBD exceeded 20% for the majority of mRNA vaccinees, but only a minority of AdV vaccinees on day 7 (Figure 3d). This coincided with substantially higher percentages of RBD^+^ plasmablasts among memory B cells for mRNA vaccinees compared to AdV vaccinees on ∼day 7 (Figure 3e). Spike^+^ plasmablast frequencies were elevated among both mRNA and AdV vaccinees on day 7 (Figure 3f).

**Table 2.** B cell frequencies by vaccine type and time-point

|  | **AdV** | **mRNA** | **p-value\*** |
| --- | --- | --- | --- |
| **Pre vaccine** | **n = 5** | **n = 20** |  |
| B cells, % of PBMC | 9.4 (7.2-11.8) | 9.0 (8.0-10.1) | 1.0 |
| B memory cells, % of B cells | 25 (21-26) | 22 (16-29) | 0.53 |
| Plasmablasts, % of B cells | 0.8 (0.4-2.8) | 0.9 (0.6-1.3) | 0.818 |
| RBD+, % of B memory cells | 0.03 (0.02-0.04) | 0.04 (0.02-0.05) | 0.518 |
| Omicron spike+, % of B memory cells | 0.10 (0.08-0.20) | 0.07 (0.06-0.09) | 0.083 |
| **Post vaccine ~ day 7** | **n = 7** | **n = 30** |  |
| Day median day (range) | 7 (5-9) | 7 (6-13) | 0.373 |
| B cells, % of PBMC | 9.2 (7.1-8.9) | 8.6 (6.3-10.8) | 0.898 |
| B memory cells, % of B cells | 27 (23-30) | 25 (18-28) | 0.443 |
| Plasmablasts, % of B cells | 2.9 (1.3-3.5) | 1.8 (1.3-2.5) | 0.565 |
| RBD+, % of B memory cells | 0.07 (0.07-0.15) | 0.58 (0.31-0.88) | 0.0001 |
| Omicron spike+, % of B memory cells | 0.18 (0.17-0.24) | 0.53 (0.41-0.98) | 0.0003 |
| **Post vaccine ~ day 14** | **n = 7** | **n = 30** |  |
| Day, median day (range) | 15 (14-20) | 16 (14-23) | 0.449 |
| B cells, % of PBMC | 10.0 (7.8-11.7) | 8.0 (6.5-9.5) | 0.356 |
| B memory cells, % of B cells | 23 (20-28) | 24 (16-27) | 0.854 |
| Plasmablasts, % of B cells | 1.4 (0.8-2.3) | 1.1 (0.7-1.5) | 0.44 |
| RBD+, % of B memory cells | 0.08 (0.06-0.18) | 0.16 (0.10-0.28) | 0.042 |
| Omicron spike+, % of B memory cells | 0.26 (0.22-0.44) | 0.35 (0.26-0.51) | 0.327 |

\*p-values for the Wilcoxon rank-sum test

The isotype distribution of vaccine-reactive B cells was similar for AdV and mRNA vaccinees (Supplementary Figure 3). Percentages of spike^+^ B cells that cross-recognized Omicron spike (34% to 41%) were also similar (Supplementary Figure 4).

### Adenovirus type 5 (Ad5) hexon protein-reactive IgG titres

Anti-Ad5 hexon IgG titres were determined by ELISA (Figure 4a) for 93 AdV vaccinees who provided pre and post-vaccination sera. Seven mRNA vaccinees were included as controls. All participants tested had detectable Ad5 hexon-reactive IgG indicating that they had prior Adenovirus infection (Figure 4b). Median rise in anti-Ad5 hexon IgG titre after AdV vaccination was 2.2-fold (interquartile range 1.5-3.1). Titres rose at least 1.5 fold for 79/93 (85%) of the AdV vaccinees but for none of the mRNA vaccinees. Four of seven participants who lacked detectable sVNT antibodies had a rise in anti-Ad5 hexon antibody titre exceeding 1.5 fold (Figure 4b, e). However, Ad5 hexon-reactive IgG titres and titre rises were not correlated with SARS-CoV-2 sVNT titres (Figure 4c-e).

**Figure 4.**
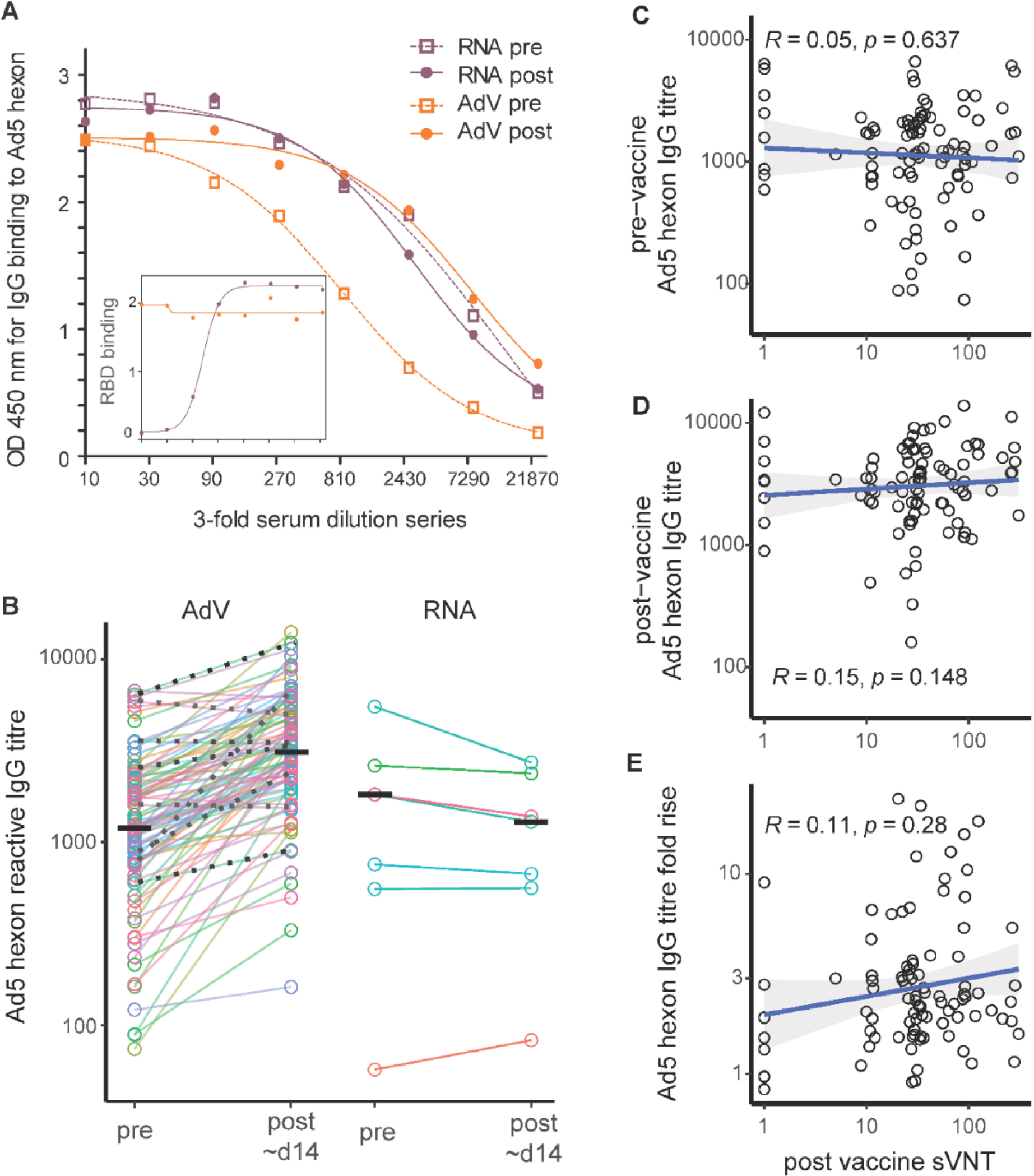
Effect of COVID-19 vaccination on antibodies against Adenovirus type 5 (Ad5) hexon. (A) Anti-Ad5 hexon IgG titration curves of pre and post vaccination sera from one AdV and one mRNA vaccinee. Inset shows post-vaccination sera from the same two individuals titrated by sVNT assay. (B) Anti-Ad5 hexon IgG titres for 96 AdV vaccinees and a subset of 7 mRNA vaccinees who had pre- and post-vaccination sera. Lines connect paired sera. Black horizontal bars indicate medians. Dotted black lines indicate AdV vaccines whose post-vaccination sVNT titres were undetectable. (C) Pre-vaccination Ad5 hexon IgG titre versus day 14 sVNT titre. (D) Post-vaccination Ad5 hexon IgG titre versus day 14 sVNT titre. (E) Pre-to-post-vaccination fold-rise in Ad5 hexon IgG titre versus day 14 sVNT titre. Each dot represents a unique sample collected from 96 AdV vaccinees. Spearman’s correlation coefficient R and associated p-values are shown.

## Discussion

The study described here provides robust evidence that that AdV AZD1222 vaccine induces substantially lower sVNT antibody titres than mRNA BNT162b2 vaccine among SARS-CoV-2 naïve HCWs. The study assessed serial dilutions of sera rather than a single serum dilution, and was large and covered a broad age range compared to previous studies ^14–18^. Estimated sVNT titres remained substantially higher among mRNA vaccinees after adjusting for age. Notably, nearly half of the AdV vaccinees lacked detectable sVNT antibodies by around 125 days after vaccination compared to only 8% of mRNA vaccinees. Factors that could contribute to differences in the immunogenicity of AdV compared to mRNA vaccines include the amount and form of protein expressed, and anti-vector immunity. To investigate the mechanism we compared spike binding antibody titres, frequencies of spike and RBD reactive B cells, and titres of IgG reactive with human Ad5 hexon.

Several results suggest that AdV vaccine may be particularly poor at inducing antibodies that target RBD. First, titres of sVNT antibodies, which inhibit RBD binding to ACE2, were around 4-fold lower among AdV compared to mRNA vaccinees whereas spike binding antibody titres were around 2-fold lower. Second, B cells that target the RBD of spike were barely detected after AdV vaccination whereas there was a small but detectable rise in spike binding B cells. This could reflect the form of protein encoded by AdV AZD1222 versus mRNA vaccine. AdV vaccine DNA uses an artificial leader sequence, which may affect the structure of the N terminus, and has to be transcribed, which can result in the production of splice variants and truncated protein^5^. Moreover, the mRNA, but not the AdV AZD1222 vaccine, encodes prefusion stabilized spike ^5^. Vaccines containing prefusion-stabilized spike have been found to induce relatively more antibody against prefusion spike S1 subunit, the target of most nAbs, whereas vaccines containing non-stabilized spike induce relatively more antibody against post-fusion spike and the S2 subunit ^22^.

Antibodies against human Ad5 hexon were detected in all participants and were boosted in most participants after receiving AdV vaccine, indicating that human adenovirus reactive memory B cells may cross-react with ChAdOx1. This is also suggested by a study that finds that plasmablast and TfH associated genes are up-regulated early after a first vaccination with AdV but not mRNA vaccine; before anti-spike antibodies are detected; and coincident with an increase in ChAdOx1 hexon reactive T cells {Ryan, 2023 #32}. However, anti-Ad5 hexon antibody titres did not correlate with post-vaccination sVNT titres, similar to other studies of AZD1222 and of recombinant human adenovirus type 26 and type 5 vaccines {Byazrova, 2022 #35}. Clinical trials indicate that ChAdOx1 neutralizing antibodies are induced by AZD12222, but titres do not correlate with anti-spike antibody titres ^2, 23^. Nevertheless, some participants had an antibody titre rise against Ad5 hexon, but not against spike, after AdV vaccination indicating that immune responses could have been biased towards the vector.

Several aspects of the study limit interpretation, in particular the timing of sample collection. Samples were mainly collected around 14 days after vaccine dose 2, but not after other doses, and the number of participants that provided blood for PBMCs on ∼ day 7 and/or 14 was small. Therefore, we could not assess the impact of anti-vector responses induced after dose 1, or correlate antibody titres with protection. In addition, we could not assess whether these vaccines induce different innate immune responses, which requires sample collection around 2 days after vaccination {Arunachalam, 2021 #33}. This is a possibility since single stranded RNA is recognized by endosomal toll-like-receptors (TLR3 and TLR7), and ChAdOx1 viral vector dsDNA is recognized by TLR9 ^24^. We also do not know how much spike protein is produced following AdV versus mRNA vaccine, and if differences in immunogenicity could be due to antigen dose. Although AdV and mRNA vaccinees were generally well matched, the interval between dose 1 and 2 was shorter for mRNA than for AdV vaccine. However, it is unlikely that a shorter interval between doses accounts for higher antibody titres, since antibody titres increased with interval between doses of mRNA vaccine. Finally, we did not report antibody titres against SARS-CoV-2 variants, we have performed sVNT with RBD of B.1.1.529 (Omicron, YP_009724390.1) and detected only low titres (≤ 20) in only 6/63 sera collected after vaccine dose 2.

Our findings that sVNT antibody titres were lower after two doses of AdV compared to mRNA vaccine are consistent with studies showing greater field effectiveness of mRNA compared to AdV vaccine ^28–30^. Subsequent studies showed that heterologous vaccination with mRNA following two doses of AdV is more effective and immunogenic than three doses of AdV vaccine, and as good three doses of mRNA^25, 26^. Consequently, WHO guidance was updated, and the majority of AdV vaccinees in our study received a 3^rd^ dose mRNA vaccine. AdV vaccines induce robust spike reactive T cell responses when used for priming ^26, 27^, which may account for their capacity to boost antibody responses to subsequent vaccination with mRNA.

In summary, this study provides further evidence that AdV AZD1222 induces low neutralizing antibody titres compared to mRNA BNT162b2 vaccine. Pre-existing vector reactive antibodies were commonly detected and boosted by AdV AZD1222 vaccine but did not appear to compromise immunity. Spike reactive B cells were induced at relatively low levels following AdV vaccine whereas RBD reactive B cells were barely detected. Together, the results suggests that differences in the immunogenicity of AdV AZD1222 versus mRNA BNT162b2 vaccines reflect the amount or form of spike protein expressed rather than anti-vector immunity. Further work is needed to improve the immunogenicity and effectiveness of viral vector vaccines as they remain an important option for pandemic and outbreak response.

## Data Availability

All data produced in the present work are contained in the manuscript.

## Acknowledgements

The study authors wish to thank the healthcare workers participating in this study. We also wish to thank other laboratory and nursing staff from each hospital who have assisted with the study: Rosemary Joyce, Meredith Krieg, Jordann Davis, Luke Blakeway, Dao Hoa Anh Huynh, Li Zhou, Kristy Nichol, Prabuddha Pathinayake, Lakshitha Gunawardhana Sara Cook, Cazz Finucane, Jaslyn Ong, Echo Wang, Ushma Wadia, and Cheryl Jones.

The WHO Collaborating Centre for Reference and Research on Influenza is funded by the Australian Government Department of Health.

This work was funded by the National Institutes of Health [R01AI141534 to SGS, AF, AJK].

## Author contributions

SGS, AF, AJK conceived the study; KS provided guidance on the design of laboratory assays; YL, LC, SS, AF, JH, YYT conducted laboratory assays; LD coordinated sites managements; CD, AK, MM, SD, MH, KER, AJ, JH, MK, SM coordinated participant recruitment and data collection; ACC, KM, AK, HM, JC, CCB, PM led the study in their respective recruitment sites; AK, DH, AF and SGS contributed to data analysis and interpretation; AF drafted the manuscript and handled all revisions; all authors contributed to interpretation of the results and development of the manuscript.

---

**Supplementary Table 1:** Characteristics of participants included in analysis of spike binding antibodies

|  | AdV | mRNA | p |
| --- | --- | --- | --- |
| **n** | 92 | 93 |  |
| **Female, n (%)** | 78 (85) | 229 (84) | 1 |
| **Age Y** | 42.5(31.2-53.0) | 37.3(31.5-45.4) | .020 |
| **Days from dose 2 to acute testing** | 15 (14-19) | 16 (14-21) | .0104 |
| **Days from dose 2 to end of season testing** | 125 (112-131) | 162 (141-200) | <.0001 |
| **Acute sVNT IC50 titre** | 31 (23-63) | 129 (85-222) | <.0001 |
| **End of season sVNT IC50 titre** | 10 (1-28) | 28 (21-32) | <.0001 |
| **Fold reduction in sVNT titre** | 4 (2.4-16.6) | 5.1 (3.2-8.7) | .855 |
| **Acute sVNT antibody undetectable, n (%)** | 5 (5) | 0 | .068 |
| **End of season sVNT antibody undetectable, n (%)** | 42 (46) | 7 (8) | <.0001 |

Continuous variables are presented as medians and interquartile ranges.

**Supplementary Figure 1.**
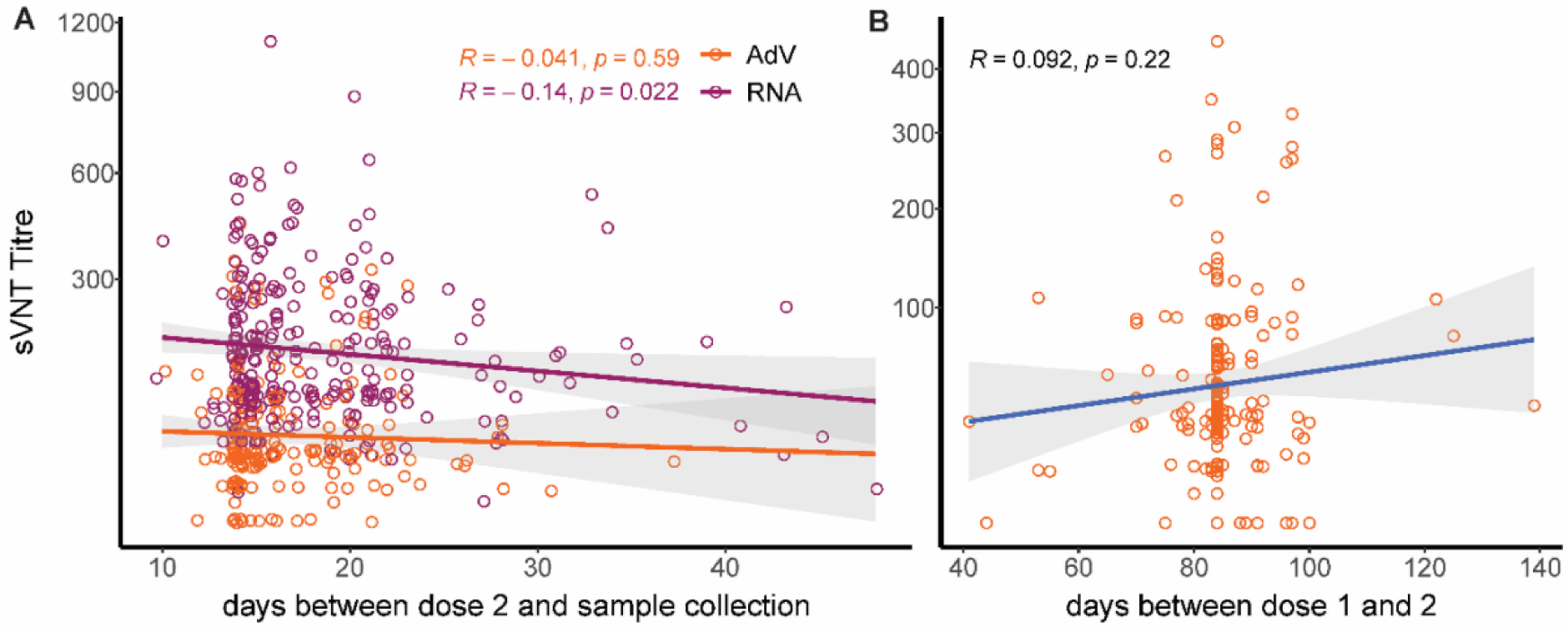
Effect of dose interval and time to sample collection on sVNT antibody titres. Data are shown with Spearman’s correlations. Each symbol represents a unique individual.

**Supplementary Figure 2.**
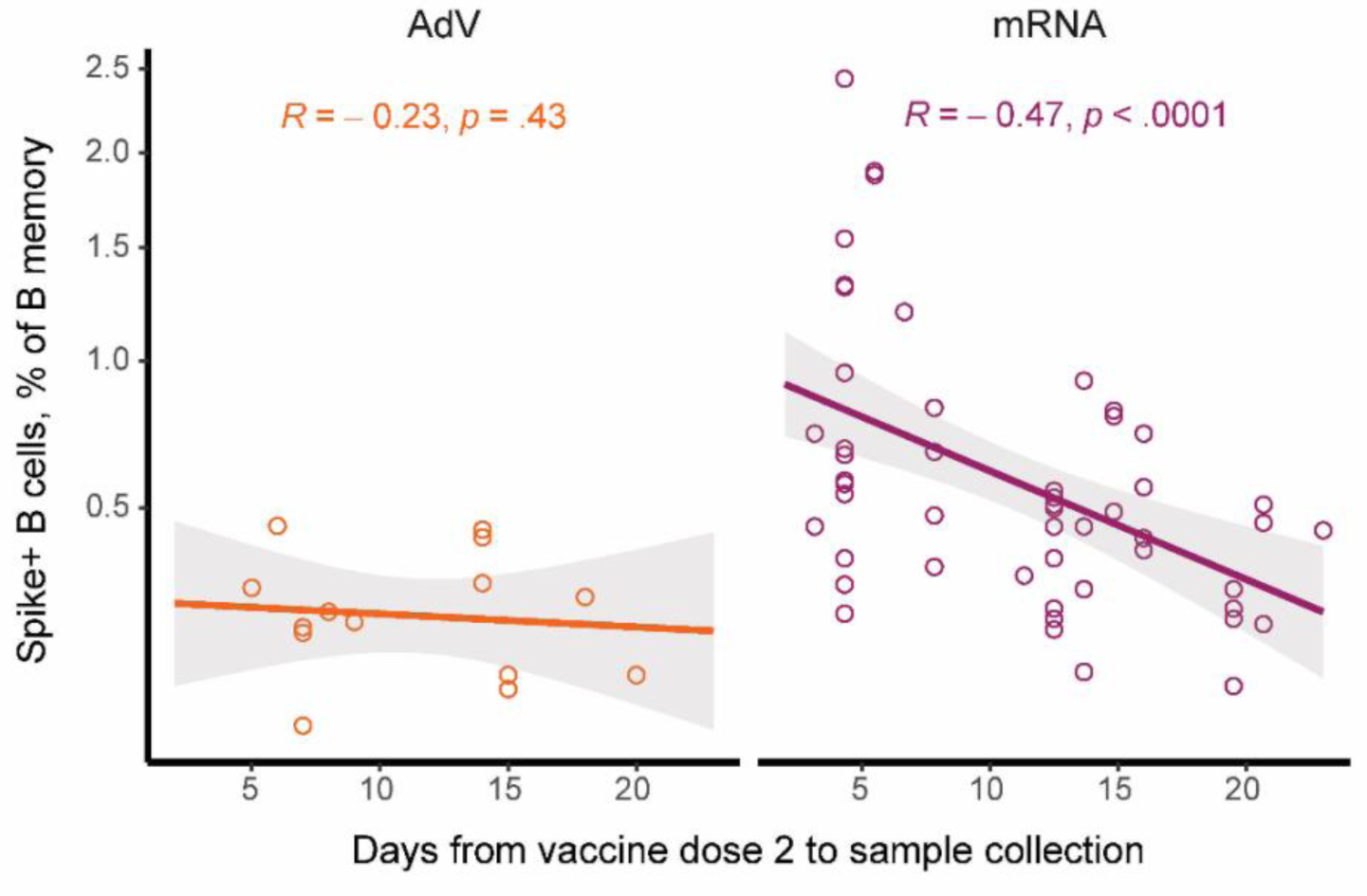
Variable effect of time to sample collection on spike+ B cell frequency. Data for both post-vaccination time-points are shown with Spearman’s correlations. Each dot represents a unique sample collected from 7 AdV vaccinees and 34 mRNA vaccinees at 1 or 2 time-points.

**Supplementary Figure 3.**
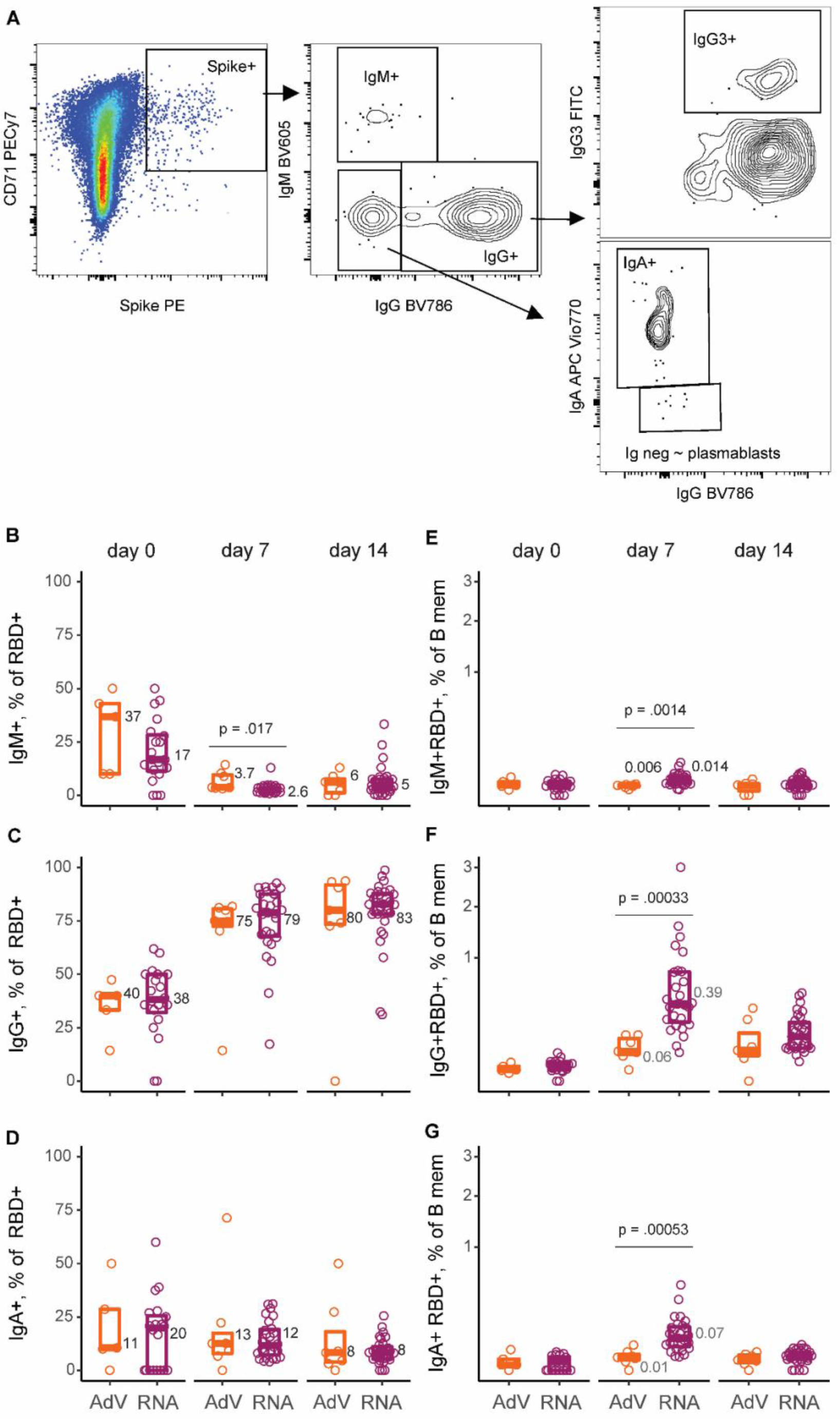
Isotype of RBD^+^ memory B cells. (A) Gates used to determine the isotype of antigen specific B cells, including IgM, IgG, IgA, or IgG3. (B-D) Percentages of RBD^+^ memory B cells with IgM, IgG, or IgA isotypes summarized for all vaccinees. (E-G) Percentages of memory B cells that are RBD^+^ and that are IgM^+^, IgG^+^ or IgA^+^ summarized for all vaccinees. Each participant is shown as a single symbol with box plots representing medians and interquartile ranges. Median values are shown to the right of each box. Significant p values for comparison of vaccine types are shown.

**Supplementary Figure 4.**
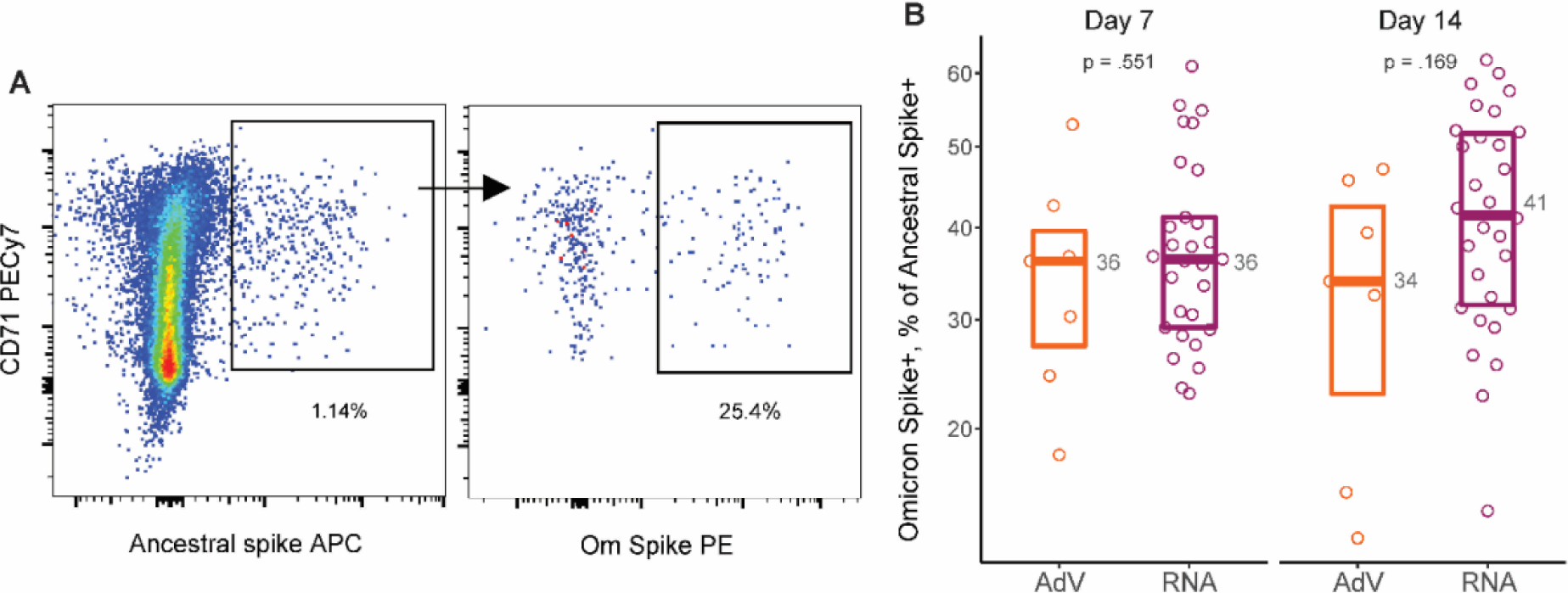
Percentage of ancestral strain spike^+^ B memory cells that cross recognize Omicron strain spike. (A) Gates used to determine the percentage of ancestral strain spike reactive B cells that cross-recognize Omicron spike (B) Results for all samples are tested are shown with boxplots representing medians and interquartile ranges. P values are shown for comparison of vaccine types. Median values are shown to the right of each box.

**Supplementary Figure 5.**
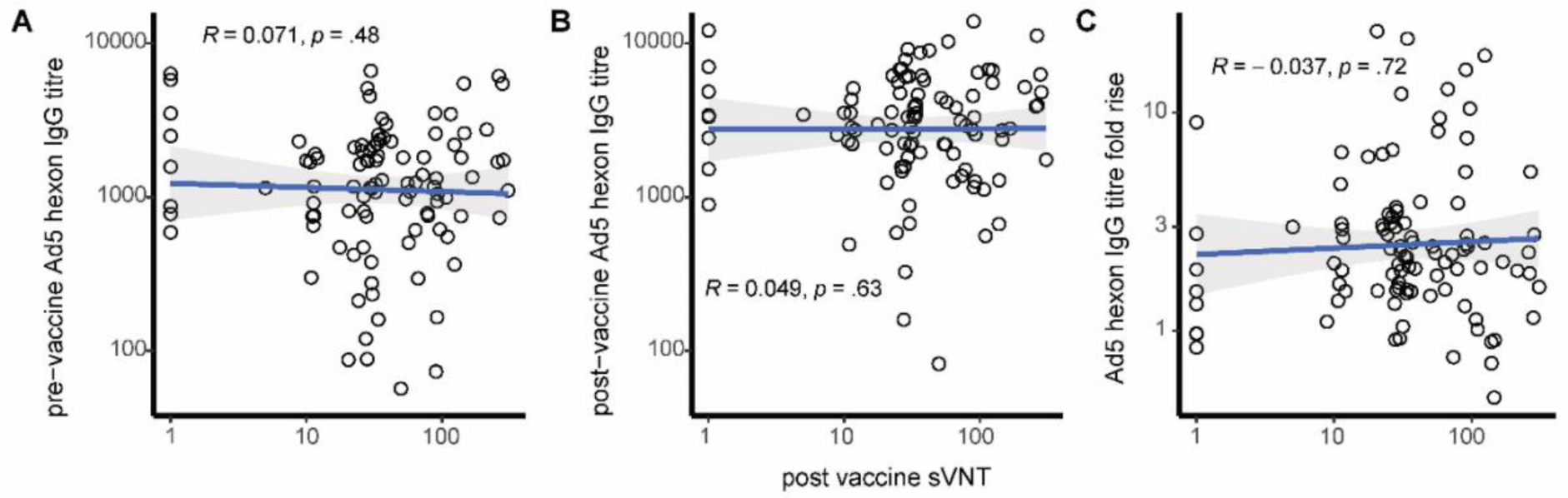
Post vaccination sVNT titres were not correlated with Ad5 hexon IgG titres. (A) Pre-vaccination Ad5 hexon IgG titre versus day 14 sVNT titre. (B) Post-vaccination Ad5 hexon IgG titre versus day 14 sVNT titre. (C) Pre to post vaccination fold-rise in Ad5 hexon IgG titre versus day 14 sVNT titre. Spearman’s correlation values are shown. Each dot represents a unique sample collected from 96 AdV vaccinees.

